# COVID-19-Associated Acute Disseminated Encephalomyelitis – A Case Report

**DOI:** 10.1101/2020.04.16.20068148

**Authors:** Tianshu Zhang, Michael B. Rodricks, Ellen Hirsh

## Abstract

We present the first described case of acute disseminated encephalomyelitis (ADEM) in a COVID-19 patient. The clinical features with head CT and brain MRI changes were described. Our described case is an atypical presentation of the novel coronavirus in a young patient and is illustrative of the possible approaches to explore the treatable differential etiologies.

## Introduction

The recent pandemic caused by the novel coronavirus (COVID-19) has been staggering in terms of the scope and speed with which the healthcare system has come under pressure. Patients typically present with fever, cough, myalgia and eventually become dyspneic. Despite the characteristic symptoms of the novel coronavirus, we have frequently noted atypical symptoms including co-infections with disparate viruses and bacteria, gastrointestinal symptoms and neurological symptoms including headache, altered mental status, loss of olfaction and seizures. We present the case of a patient with significant neurological symptoms.

## Case

A right handed female in her early forties presented to the emergency department after she developed dysphagia, dysarthria and encephalopathy two days prior to admission. She had a history of well controlled hypertension and dyslipidemia. The patient had initially become ill eleven days prior to admission with a headache and myalgia. She was seen by her primary care physician and treated as an outpatient with a course of azithromycin. She had a negative influenza swab and a negative rapid strep test at the office. She was, however, not checked for COVID-19 due to the lack of testing capacity. The patient’s relative, with whom she resided, had recently returned from a trip aboard. The day after returning her relative developed a headache and myalgia which were mild, lasted four days and were self-limited. Her relative did not seek any medical care. The patient fell ill four days after her relative’s return with similar symptoms of a headache and myalgia.

In the emergency department the patient was noted to have a temperature of 102.2 degrees Fahrenheit with some mild tachypnea and hypoxemia. The physical examination revealed a patient in no acute distress with diffuse rhonchi on auscultation. Neurologically she was awake and alert and followed commands although sluggishly. She had difficulty with her speech with components of both dysarthria and expressive aphasia. Additionally there was bulbar impairment with difficulty handling secretions and dysphagia. There was no meningismus which could be elicited. Her pupils were equal round and reactive but she showed a right gaze preference and a mild left facial droop. She had mildly decreased equal bilateral strength. Her deep tendon reflexes were preserved.

Initial laboratory studies identified a mild leukocytosis with lymphopenia. The chest x-ray showed patchy consolidation in the right lower lung. A non-contrast computed tomography (CT) of the head showed no evidence of intracranial hemorrhage, but there were multifocal patchy areas of white matter hypoattenuation (Fig. 1). A lumbar puncture was performed to clarify the diagnosis and to exclude central nervous system infection. Cerebrospinal fluid (CSF) analysis revealed normal cell counts, protein and glucose. A polymerase chain reaction (PCR) panel for meningitis and encephalitis, including herpes simplex 1 and 2, human herpes 6, Cryptococcus and Varicella-Zoster virus was entirely negative as were bacterial cultures. An electroencephalogram (EEG) did not show electrical evidence of seizures. The COVID-19 test became available 2 days after admission and detected the novel coronavirus (SARS-CoV-2) target nucleic acid.

**Fig. 1.**
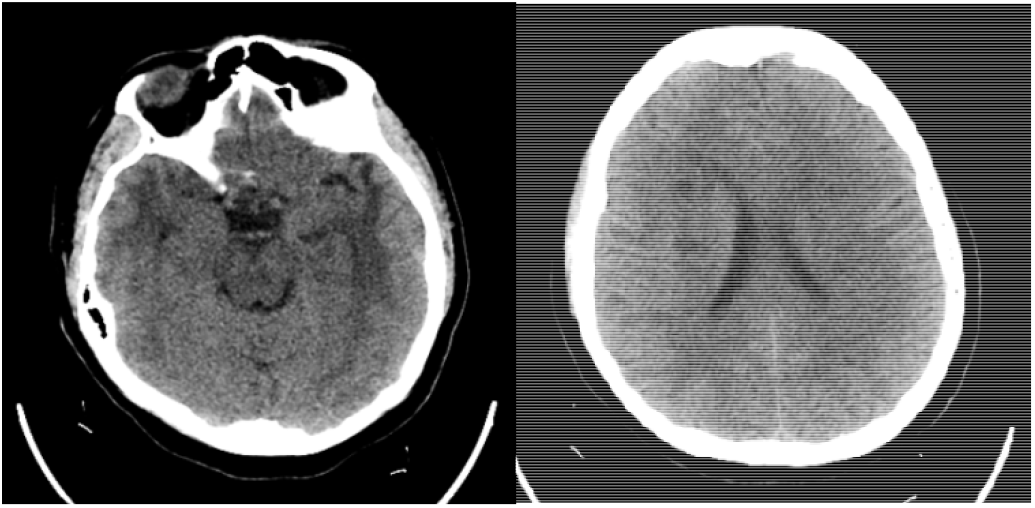
CT head - bilateral temporal and subcortical hypodensity

To assess the white matter lesions found on head CT, a magnetic resonance imaging (MRI) of the brain with and without contrast was obtained. The MRI showed extensive patchy areas of abnormal signal involving bilateral frontoparietal white matter, anterior temporal lobes, basal ganglia, external capsules and thalami. Additionally some of these foci demonstrated diffusion weighted imaging (DWI) changes and corresponding apparent diffusion coefficient (ADC) changes, with questionable minimal enhancement (Fig. 2, Fig. 3). Magnetic resonance angiography (MRA) of the brain and neck were essentially normal.

**Fig. 2.**
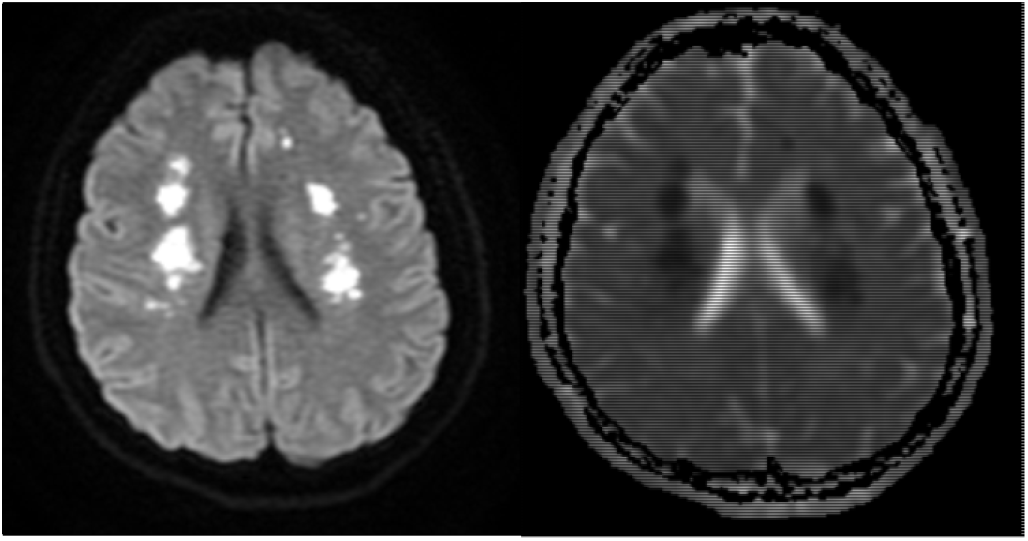
MRI brain - subcortical diffusion weighted imaging (DWI) and apparent diffusion coefficient (ADC) map changes

**Fig. 3.**
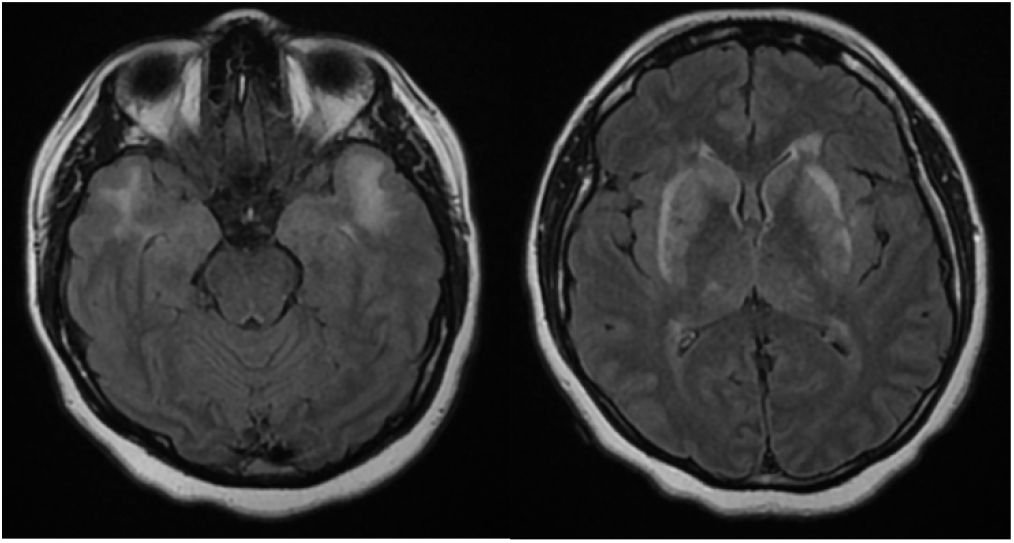
MRI brain - subcortical and temporal T2 FLAIR changes

## Discussion

It is important to consider other infectious etiologies with or without a diagnosis of COVID-19 in a patient with fever, neurological deficits and encephalopathy. An electroencephalogram (EEG) is indicated to exclude subclinical seizures and non-convulsive status epilepticus. The CT findings prompted a further work up for cerebral abscesses and septic emboli. It is difficult logistically to perform an MRI on a COVID-19 positive patient or a person under investigation (PUI) due to the concern for contamination of equipment, the requisite patient transportation and the risk of exposure to healthcare providers. Given the clinical picture and the head CT findings; however, it seemed rational to take the necessary steps to obtain an MRI. The brain MRI showed multiple DWI lesions and corresponding ADC sequence changes which were most consistent with hypoxic-ischemic injury from systemic perturbations and resultant tissue hypoxia. The predominance of the extensive T2 FLAIR signal changes in the cerebral white matter lesions is more suggestive of a demyelinating process. These lesions are extensive, bilateral, and predominantly subcortical, with additional involvement of the deep nuclei. These features are most compatible with the typical description of acute disseminated encephalomyelitis (ADEM), especially in the setting of a recent or ongoing viral infection with acute neurologic deficits. Multiple sclerosis is within the differential diagnosis but given the clinical picture described as well as the diagnosis of COVID-19 it seems a very unlikely etiology of the clinical picture.

The patient was treated with hydroxychloroquine, ceftriaxone and a five day course of intravenous immunoglobulins (IVIG). Steroids were not used as it was felt as possibly detrimental given the acute COVID-19 diagnosis and in keeping with current guidelines. After five days of IVIG the patient was showing signs of improvement-better able to handle secretions, less dysarthria and afebrile with no respiratory symptoms.

## Conclusion

We present the first described case of ADEM in a COVID-19 patient. ADEM is an acute inflammatory demyelinating condition of the central nervous system that rarely occurs after a viral infection. ADEM has been described in cases of the Middle East respiratory syndrome coronas virus (MERS) (2) and Coronavirus OC43 (3). A recent case reported acute hemorrhagic necrotizing encephalopathy in a COVID-19 patient as a rare super acute attack of immune-mediated CNS injury with unique CT and MRI features (1). Our described case is an atypical presentation of the novel coronavirus in a young patient and is illustrative of the possible approaches to explore the treatable differential etiologies.

## Data Availability

Contact to the authors -

